# Cohort profile: the ‘Biomarkers of heterogeneity in type 1 diabetes’ study – a prospective cohort study of clinical and metabolic phenotyping of individuals with long-standing type 1 diabetes

**DOI:** 10.1101/2023.08.17.23294197

**Authors:** Dick Mul, Rita D.M. Varkevisser, Henk-Jan Aanstoot, Pim Dekker, Erwin Birnie, Erik Boersma, Lianne S.M. Boesten, Michael P. Brugts, Peter R. van Dijk, Petronella H.L.M. Duijvestijn, Sanjoy Dutta, Christine Fransman, Rob K. Gonera, Klaas Hoogenberg, Adriaan Kooy, Esther Latres, Sandra Loves, Giesje Nefs, Theo Sas, Frederik A.J. Verburg, Charlotte E. Vollenbrock, Marleen J. Vosjan-Noeverman, Martine M.C. de Vries-Velraeds, Henk J. Veeze, Bruce H.R. Wolffenbuttel, Melanie M. van der Klauw

**Author notes:** **Correspondence to:** Pim Dekker.

## Abstract

**Purpose:** The ‘Biomarkers of heterogeneity in type 1 diabetes’ study cohort was set up to identify genetic, physiological and psychosocial factors explaining the observed heterogeneity in disease progression and the development of complications in people with long-standing type 1 diabetes (T1D). Data and samples are available for new studies and collaborations.

**Participants:** Data- and samples were collected in two subsets. 1) A prospective cohort of 611 participants aged ≥16 years with ≥5 years T1D duration was recruited from four Dutch Diabetes clinics between June 2016 and March 2021. At baseline and 1- and 2-year follow-up visits, physical assessments were performed, and blood and urine samples were collected. Participants completed questionnaires about diabetes-related problems, quality of life, neuropathy and impaired awareness of hypoglycaemia at baseline and at the last follow-up visit. A subgroup of participants underwent mixed-meal tolerance tests (MMTT) at baseline (n=169) and at 1-year follow-up (n=104). Genetic data and linkage to medical and administrative records were also available. 2) A second cross-sectional cohort, aiming to include 200 participants aged ≥18 years with ≥35 years T1D duration, was recruited from 7 centres, collecting measurements and samples plus 5-year retrospective data.

**Findings to date:** Fasting residual C-peptide secretion associated with decreased risk of impaired awareness of hypoglycaemia. Stimulated residual C-peptide was detectable in an additional 10% of individuals compared with fasting residual C-peptide secretion. MMTT measurements at 90 minutes and 120 minutes showed good concordance with the MMTT total area under the curve. An overall decrease of C-peptide at 1-year follow-up was observed.

**Future plans:** Research groups are invited to consider the use of this data and sample collection. Future work will include additional hormones, beta-cell-directed autoimmunity, specific immune markers, microRNA’s, metabolomics and gene expression data, combined with glucometrics, anthropometric/clinical data and additional markers of residual beta-cell function.

**Strengths and limitations of this study:** *Strengths:*

- The Biomarker cohort is a large longitudinal prospective cohort study with three time points, collecting biosamples and clinical data from participants with well-established and long-standing type 1 diabetes (≥5 years).
- A subgroup with detailed clinical data underwent MMTT tests at two timepoints allowing further residual beta-cell marker studies.
- The Biomarker and Long-Term type 1 Diabetes cohorts represent a “real-world” population, also including participants from non-academic/-specialised centres.

*Limitations:*

- Despite the fact that data and biosamples were collected from more than 600 participants, this number may be too low for (sub) stratification of the data (e.g. insulin delivery modality, different treating centres and therapies etc.).
- In the prospective group there was a relatively high dropout rate of 25% after 2 years, largely affected by the Covid-19 outbreak.

## INTRODUCTION

Type 1 diabetes (T1D) is characterized by severe insulin deficiency caused by insulin-producing beta-cell dysfunction or and other defects that result in impaired insulin production. This may induce secondary autoimmune responses that damage beta-cells and disturb their coherence and function in the islets of Langerhans.^1 2^ This disease process occurs in people with specific genetic backgrounds and is characterized by two pre-symptomatic phases (stage 1 and 2). Stage 3 is the phase of appearance of clinical symptoms, establishment of the final diagnosis and initiation of treatment. ^3^ Stage 4 is long-standing T1D.^4^

Progression from stage 1/2 to clinical diagnosis includes both immune dysregulation and beta-cell dysfunction. Several phase 2 clinical studies involving immunomodulation have demonstrated benefits in both stage 2 and 3 T1D. ^5^ Stage 3 and 4 T1D (long-standing T1D ^4^) are characterized by progressive loss of beta-cell mass, resulting in the need for life-long insulin suppletion to survive. However, residual C-peptide production has been demonstrated in long-standing diabetes, ^1 6^ conferring clinical benefits. Despite the currently available treatments and technology, reaching glycaemic targets is very difficult if not impossible for most Persons With T1D (PWDs). ^7^ Consequently exposure to suboptimal glucose levels causes micro- and macrovascular, psychological and psychosocial complications in the long run in stage 4. These complications are strongly dependent on the PWD’s historical glycaemic regulation, which in turn is determined by many factors and shows extensive heterogeneity between PWDs. These factors include biological factors (i.e. genetic predisposition to insulin resistance and/or vascular damage, residual insulin production), ^8^ the availability and access to healthcare, medication and technology, ^9^ and psychosocial factors.^10^

Despite medical and technological progress, T1D still has a profound impact on life-expectancy, leading to a loss of 10 to 18 life years, depending on age of diagnosis. ^11^ To prevent the development of T1D-related complications, international guidelines recommend HbA1c levels below 7% (53 mmol/mol)^12^ or time in blood glucose range (TIR) > 70%. ^13^ However, only a minority of PWDs are currently achieving the recommended HbA1c levels. ^7 14^ This may partly be explained by extensive heterogeneity in pathology seen in all four stages of T1D. Clinical symptoms and severe metabolic disturbance (e.g. diabetic ketoacidosis ^15^) at onset, autoimmune markers before and after onset, initial glycaemic outcomes (HbA1c, acute glucose levels, TIR), and the efficacy of therapeutic interventions^8 16^ all vary between PWDs. Furthermore, the T1D phenotype represents different distinct underlying functional or pathobiological mechanisms, also called endotypes. ^17^ Improving future outcomes will depend on the ability to further unravel this heterogeneity, dissect endotypes and develop individualized prediction, prevention and intervention strategies to prevent progression to stage 3 and 4 T1D and eventually even restore immunological tolerance and beta-cell mass. ^18^ Using combinations of (epi)genetic data (e.g. genetic risk loci) and disease biomarkers (clinical data, metabolic markers and immunological markers) can provide a new integrative approach that will help to develop personalized T1D interventions. ^2^

Several studies on factors of heterogeneity in T1D stages 1-3 have been initiated.^16^ However, unravelling the heterogeneity in stage 4 T1D requires appropriate clinical datasets on the individual history of the PWD and longitudinally collected samples. Together with the JDRF and the Dutch Diabetes Research Foundation (‘Diabetes Fonds’ [DF] in Dutch) we identified needs and research questions for a new project, based on research gap analysis and existing knowledge of the field. With funding from JDRF and DF we developed the ‘Biomarkers of heterogeneity in type 1 diabetes’ prospective cohort study to collect data and bio-samples from PWDs with at least 5 years diabetes duration, and the ‘Long-Term type 1 Diabetes’ cross-sectional cohort to collect data and bio-samples from PWDs with at least 35 years diabetes duration. This study ran from 2016 to end 2023. We collected clinical data and samples for physiological and genetic analyses and information on psychosocial factors, aiding in explaining the observed heterogeneity in T1D and its progression. While we have started to use data and samples from this Biomarker cohort, ^19–21^ the data and samples are available for additional projects and collaborative research. Requests for data and/or samples will be considered by a steering committee. Here we provide an overview of the study setup, a description of the participants and future aims.

### Cohort description

The prospective ‘Biomarkers of heterogeneity in type 1 diabetes’ study (Clinicaltrials.gov/ NCT04977635; called ‘Biomarker study’ from hereon) was initiated by Diabeter Netherlands (Rotterdam, The Netherlands) and the University Medical Center Groningen (Groningen, The Netherlands). These centres provided the majority of patients, limiting variation in treatment of participants. Additionally, PWDs from Haaglanden Medical Center (The Hague, The Netherlands) and Ikazia hospital (Rotterdam, The Netherlands) were recruited. The project and amendments for additional research and future research questions were approved by the Medical Ethics Review Board of the University Medical Center Groningen (METC 2015/493).

A total of around N=600 participants was deemed sufficient to stratify for age and diabetes duration. Between June 2016 and March 2021, a total of 611 people with T1D aged ≥16 years and with a diabetes duration ≥ 5 years (called the ‘Biomarker complete study cohort’ from hereon) were included (Figure 1). The T1D diagnosis was determined by either the presence of diabetes autoantibodies, based on clinical and historical data or both. Exclusion criteria were: all types of diabetes which are not considered T1D according to ADA criteria, ^22^ pregnancy (until 3 months after childbirth) and breastfeeding (until 3 months after breastfeeding), using experimental medication or participating in other studies with conflicting goals and schedules, decision against participation at the investigator’s/physician’s discretion and being unwilling to be informed on incidental findings. A subgroup of 169 participants (28%) positively responded to the invitation to participate in additional mixed meal tolerance tests (MMTT: called the ‘Biomarker MMTT-subcohort’ from hereon).

**Figure 1.**
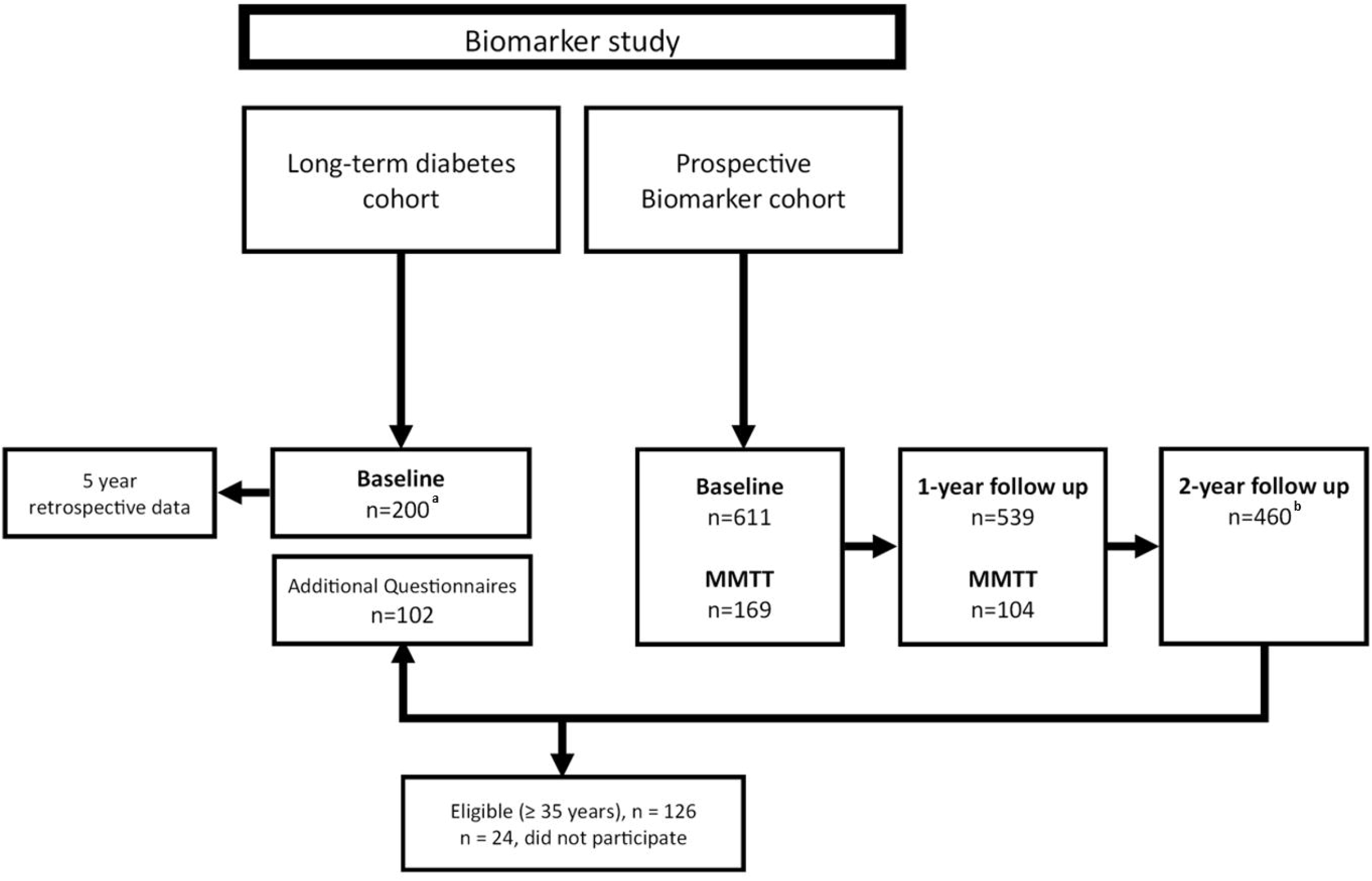
Flowchart of participant inclusion. ^a^ aim to include 200 individuals. ^b^ n=23 of these n=460 skipped the 1-year follow-up.

Participation in this sub-study was voluntary and no additional in- and exclusion criteria applied. The ’Long-Term type 1 Diabetes’ cross-sectional study (NL62401.042.17; called the ‘LTD study’ from hereon) was also initiated by the University Medical Center Groningen (Groningen, the Netherlands) and Diabeter Netherlands (Rotterdam, The Netherlands). This study aims to recruit 200 people with T1D aged ≥16 years and with a diabetes duration ≥ 35 years (‘LTD cohort’). T1D diagnosis and exclusion criteria are equivalent to the prospective Biomarker study. The cohort comprises participants from the Biomarker complete study cohort and PWDs recruited from the Martini Hospital (Groningen, the Netherlands), the Wilhelmina Hospital (Assen, the Netherlands), the Treant Hospital Group (locations Emmen, Hoogeveen and Stadskanaal, the Netherlands), the Medical Center Leeuwarden (Leeuwarden, the Netherlands), and the Dijklander Hospital (locations Hoorn and Purmerend, the Netherlands). The LTD project and study protocol amendments were approved by the Medical Ethics Review Board of the University Medical Center Groningen (METC 2017/412). Recruitment started in 2019 and is expected to close by the end of 2023.

## PARTICIPANT RECRUITMENT

Eligible participants received (electronic) flyers and notifications about the studies and were asked by their diabetes care providers about their interest to participate. Interested PWDs were contacted, provided with information and were given the opportunity to ask questions to an independent physician. After written informed consent was provided, the participant was enrolled in the study and assigned a unique study number.

### Participant and public involvement and data dissemination

Participants, funders or the public were not involved in the design, conduct, reporting or dissemination plans of this study. Participants were updated via e-mail newsletters, the Diabeter Netherlands website (www.diabeter.nl) and on social media. We have presented initial results and will present future results of the studies at national and international conferences, in peer-reviewed research papers, various other channels including local and social media, and via the research website: www.diabeterresearch.com.

## DATA COLLECTION

Table 1 lists the parameters collected from the study participants.

**Table 1:**
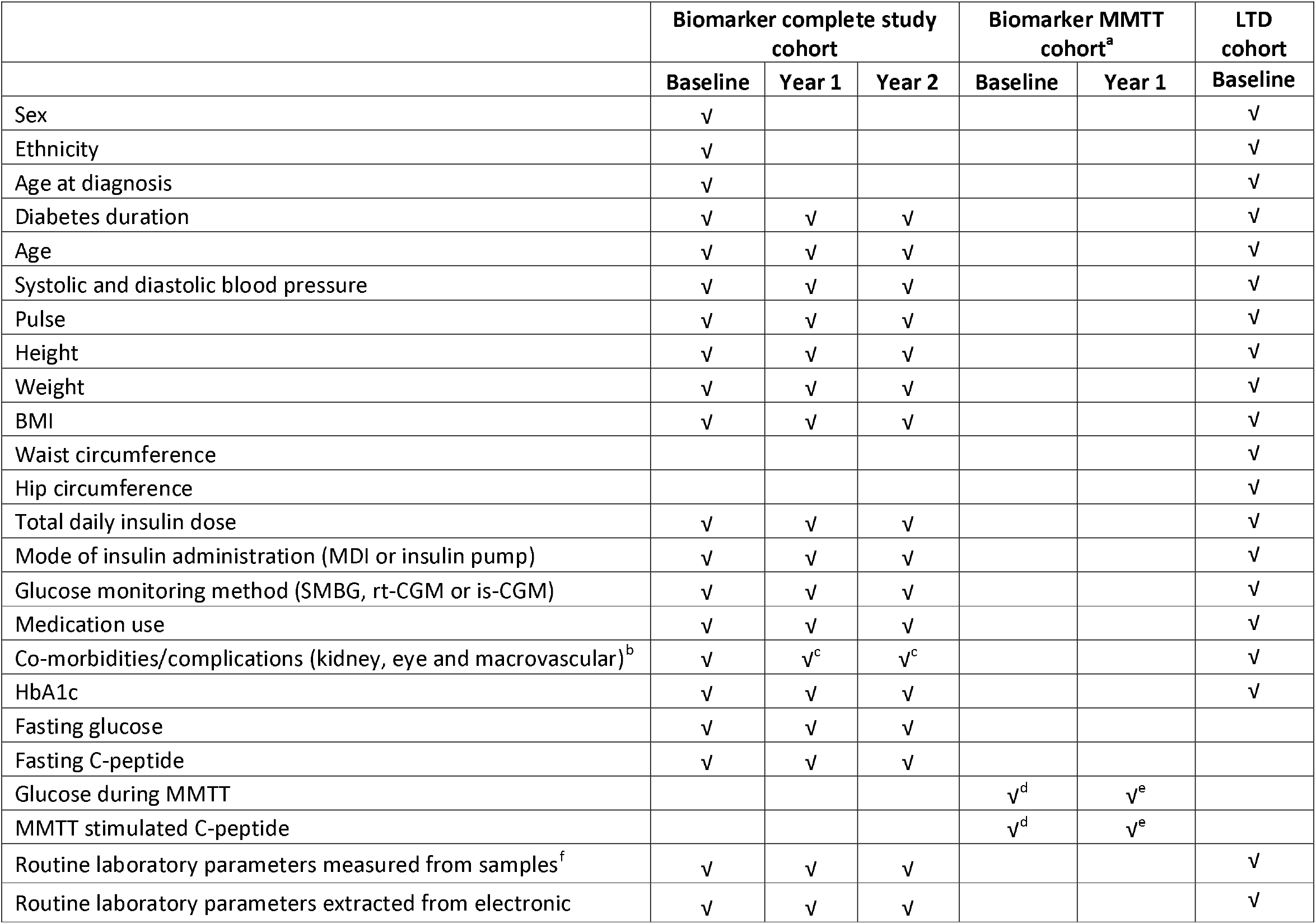
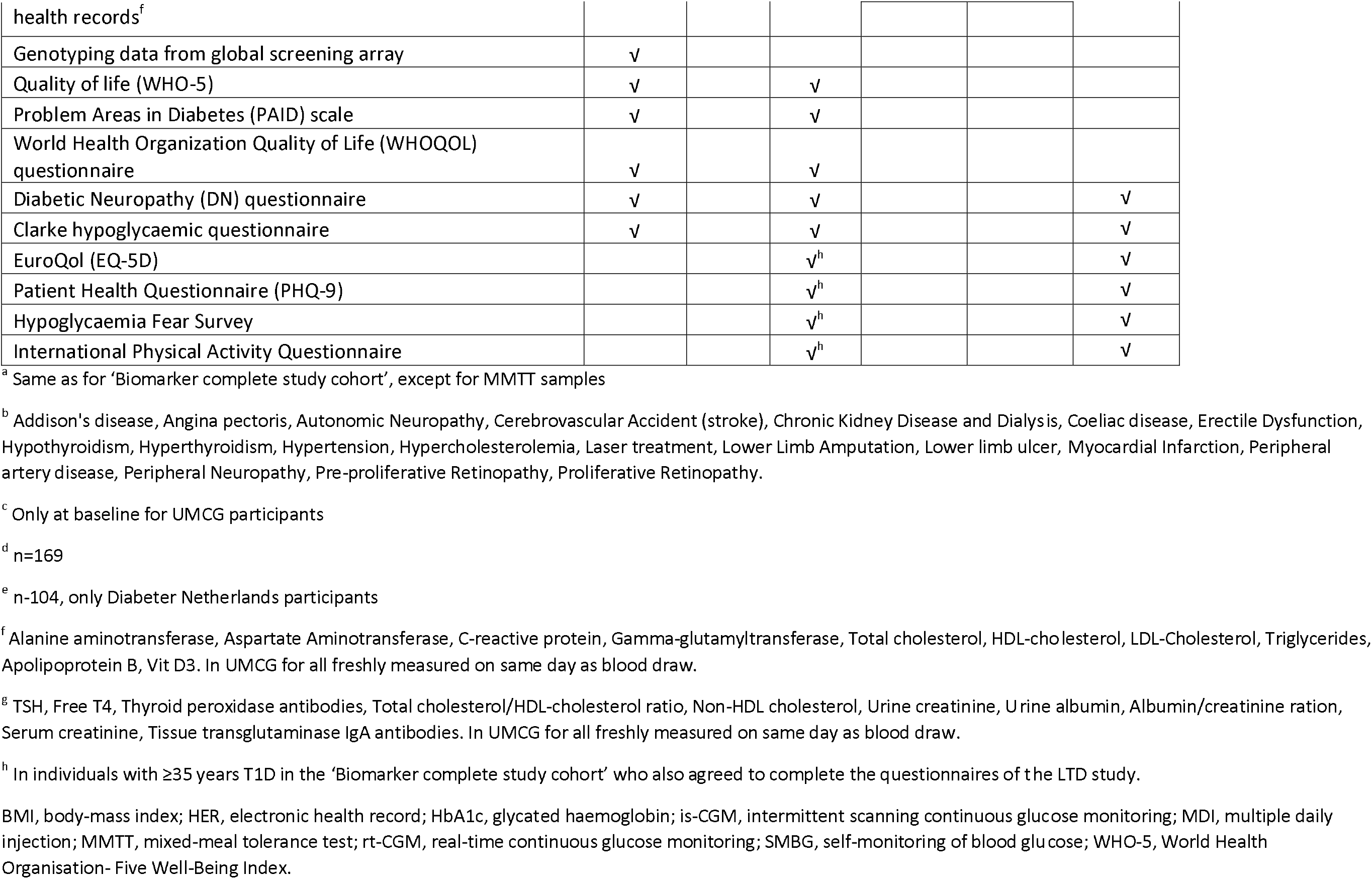
Summary of parameters collected from the cohort.

### Biomarker complete study cohort (prospective)

At the baseline visit, fasting blood and urine samples were collected. Part of the samples underwent immediate analysis of routine haematology and biochemistry, and the remainder of the samples were stored for future analysis. All participants completed six questionnaires: the World Health Organisation-Five Well-Being Index (WHO-5), ^23^ the Problem Areas In Diabetes (PAID) Scale, ^24 25^ the World Health Organization Quality of Life (WHOQOL) questionnaire, ^26^ the Diabetic Neuropathy (DN4) questionnaire^27^ and the Dutch version of the Clarke hypoglycaemic (impaired awareness of hypoglycaemia [IAH]) questionnaire, ^28 29^ either on paper or online. UMCG participants underwent anthropometric assessments and foot examination including arterial pulsation, tuning fork and monofilament evaluation^30^ by a trained physician assistant. For the PWDs attending the other participating clinics, anthropometric data were retrieved from their electronic health records by the study team after enrolment. All study procedures were repeated at the 1-year and 2-year follow-up visits, except for the questionnaires, which were repeated only at 2-year follow-up.

### Biomarker MMTT-subcohort (prospective)

Participants provided additional informed consent to participate in the MMTTs. They underwent an MMTT at baseline (n=169) and 1-year follow-up (n=104). If possible, the MMTT was carried out during the same appointment as the fasting blood and urine samples collection. Alternatively, a separate appointment was made by the study team.

### LTD cohort (cross-sectional)

At the study visit, fasting blood (when possible) and urine samples were collected. Part of the samples underwent immediate analysis of routine haematology and biochemistry and the remainder of the samples were stored for future analysis. Each participant completed paper-and-pencil questionnaires. Five questionnaires, on quality of life (EuroQol-5D [EQ-5D] ^31^) and Patient Health Questionnaire-9[PHQ-9]^32^), psychosocial burden and fears (Hypoglycaemia Fear Survey-II [HFS-II ^33^), neuropathy (DN4^27^), impaired awareness of hypoglycaemia (Dutch version of the Clarke questionnaire^28 29^), and physical activity (International Physical Activity Questionnaire [IPAQ] ^34^), were completed. Participants from all sites underwent anthropometric assessments and foot examination including arterial pulsation, tuning fork and monofilament evaluation ^30^ by a trained physician. Retrospective data up till five years before inclusion were extracted from their electronic health records by the study team after enrolment. Individuals included in the Biomarker complete study cohort with a diabetes duration ≥35years were contacted after the 2-year follow-up to fill out additional questionnaires (EQ-5D, PHQ-9, HFS-II, and IPAQ) in order to harmonise the data between the LTD cohort participants derived from the Biomarker complete study cohort. Participants were only contacted if they had previously consented to being contacted for follow-up studies. When informed consent had been provided, the additional questionnaires were sent out.

All study materials (e.g. collected samples, paper questionnaires) and study data(sets), including medical record data, results from the questionnaires and sample analysis were stored under the participant’s unique study number.

### Biochemical analyses and storage of samples and data

Biomarker complete study cohort: At each visit, blood samples were collected in coagulation (Becton-Dickinson, cat. no. 367953), lithium-heparin (Becton-Dickinson, cat. no. 367378), EDTA (Becton-Dickinson, cat. no. 367525), EDTA P800 (BD 366421) and PAXgene RNA (Qiagen, cat. no. 762165; at baseline only) blood collection tubes. PAXgene DNA tubes (Qiagen, cat. no. 761115) were collected for non-UMCG participants at one visit during the study. For UMCG participants DNA was isolated from EDTA and EDTA-P800 pellets. Blood collected in coagulation tubes was allowed to coagulate for 30 minutes. Coagulation, lithium-heparin, EDTA, EDTA P800 and urine tubes were centrifuged at room temperature for 10 minutes at 1500 rpm. Fasting morning urine was collected in tubes without any additives (Becton-Dickinson, cat. no. 365000). Blood and urine samples were aliquoted and stored at -80°C in dedicated freezers located in the clinical laboratory of the IJsselland Hospital (Capelle aan den IJssel, The Netherlands). DNA samples were genotyped using the Infinium Global Screening Array (GSA)-24 v1 and v3 Illumina Inc. (San Diego, United States).

For the MMTT procedure the target glucose level at the start of the test was between 3.3 and 12 mmol/l. If values were lower than 3.3 mmol/l, oral glucose was administered and glucose levels were checked every 30 minutes until levels were in range again. If values were higher, a correction bolus was needed, and participants were retested after 30-45 minutes. If glucose values during the night prior to the test were continuously >12 mmol/mL, tests were rescheduled. An IV-catheter was placed in a cubital vein or in the hand-wrist area from which blood was sampled. At time point 0, participants were given a dose of Resource® Protein (Nestlé) mixed meal (@ 1,25 kcal/mL), comprising 6 ml/kg body weight to a maximum of 60 kg (=360 ml). The dose had to be consumed in no more than five minutes. Blood was collected at 0, 30, 60, 90, 120 minutes in three types of tubes: coagulation tubes (Becton-Dickinson, cat. no. 367955), lithium-heparin tubes (Becton-Dickinson, cat. no. 367376) and EDTA P800 tubes (BD 366421), aliquoted and stored at -80 °C in dedicated freezers located in the clinical laboratory of the IJsselland Hospital (Capelle aan den IJssel, The Netherlands). At each time point, glucose from the drawn blood was also measured using a point-of-care glucose meter.

LTD cohort: At baseline visit, blood samples were collected in coagulation (Becton-Dickinson, cat. no. 367953), lithium-heparin (Becton-Dickinson, cat. no. 367378), EDTA (Becton-Dickinson, cat. no. 367525), trisodium citrate (Bectron-Dickinson, cat. No. 366575) and PAXgene RNA (Qiagen, cat. no. 762165) blood collection tubes. Fasting morning urine and 2-hour post-prandial urine were collected in tubes without any additives (Becton-Dickinson, cat. no. 365000). Blood collected in coagulation tubes was allowed to coagulate for 30 minutes. Coagulation, lithium-heparin, and EDTA tubes were centrifuged at room temperature for 10 minutes at 1300 rpm. DNA was isolated from EDTA pellets. Trisodium citrate tubes were centrifuged for 20 minutes at 1300 rpm. Urine sample tubes were centrifuged at 4 degrees for 10 minutes at 2000 rpm. Blood and urine samples were aliquoted and stored at -80°C in dedicated freezers at the University Medical Center Groningen.

Availability of biosamples is listed in Table 2. Data from biochemical analyses and extracted from electronic heath records were collected in multiple databases stored on secure servers at the University Medical Center Groningen and at Diabeter.

**Table 2:** Bio sample availability for baseline and 1-year and 2-year follow-ups.

| **Biosamples** | **Biosamples Baseline** | **First follow-up**<br>**(T1)** | **Second follow-up**<br>**(T2)** |
| --- | --- | --- | --- |
| *Biomarker complete study cohort* |  |  |  |
| Serum | √ | √ | √ |
| Lithium-heparin plasma | √ | √ | √ |
| EDTA (plasma) | √ | √ | √ |
| EDTA-P800 (plasma) | √ | √ | √ |
| RNA (whole blood) | √ |  |  |
| DNA (from whole blood/buffy coat) | √ |  |  |
| Urine (normal) | √ | √ | √ |
| *MMTT cohort (additional samples to biosamples Biomarker complete study cohort)* |  |  |  |
| Serum (timepoints 0, 30, 60, 90 and 120 minutes) | √ | √ |  |
| Lithium-heparin plasma (timepoints 0, 30, 60, 90 and 120 minutes) | √ | √ |  |
| EDTA-P800 plasma (timepoints 0, 30, 60, 90 and 120 minutes) | √ | √ |  |
| *LTD cohort* |  |  |  |
| Lithium-heparin | √ |  |  |
| DNA (from whole blood/buffy coat) | √ |  |  |
| Serum | √ |  |  |
| Morning urine | √ |  |  |
| Urine 2 hours post-prandial | √ |  |  |
| EDTA (plasma) | √ |  |  |
| Citrate (plasma) | √ |  |  |
| RNA (whole blood) | √ |  |  |

## PARTICIPANT CHARACTERISTICS

Table 3 shows the baseline characteristics of the study participants. The median (interquartile range [IQR]) age of the Biomarker complete study cohort participants at baseline was 31.7 (23.2–52.4) years and 59% were women. The majority of participants were of western European origin. The median age at diagnosis was 12.3 (7.9–20.6) years and median duration of diabetes was 18.5 (11.8– 29.9) years. Participants used a median total daily dose of insulin of 50 (40–64) U/day and had a mean (SD) HbA1c of 61 (13) mmol/mol (7.7 [1.1]%). The majority of participants (74%) did not produce any detectable fasting levels of C-peptide (<3.8 pmol/L). The 26% of participants who did, showed a median fasting C-peptide level of 30 (10–110) pmol/L. With most participants (61%) using an insulin pump for insulin administration, this cohort may not be representative for the general T1D population in the Netherlands. Just over half of included participants (54%) were from Diabeter Netherlands (Figures 2 and 3). Since this is a specialist centre for people with T1D, historically focusing on young children, adolescents and young adults with T1D and on technology, participants were younger (mean [SD] age: Diabeter Netherlands, 27 [10] years: other hospitals, 51 [15] years). Also the proportion of insulin pump users was higher (Diabeter Netherlands, 70%; other hospitals, 50%). It must also be noted that glucose monitoring was mostly performed by SMBG (77%), while real-time (rt-)CGM and intermittent scanning (is-)CGM were used by only 19% and 5% of participants, respectively. Rt-CGM and is-CGM use increased in the course of the study as insurance coverage of is-CGM was introduced after start of the study. Approximately one fifth of participants were using antihypertensive and lipid-lowering drugs.

**Table 3:** Participants characteristics at baseline (n=611)

|  | Biomarker complete study cohort |  | MMTT-cohort |  | LTD cohort |  |
| --- | --- | --- | --- | --- | --- | --- |
|  | Mean (SD), range (unless stated otherwise) | n | Mean (SD), range (unless stated otherwise) | n | Mean (SD), range (unless stated otherwise) | N<sup>a</sup> |
| Age, median (IQR), range (years) | 31.7 (23.2–52.4), 16.0–80.4 | 611 | 26.3 (21.5–47.9), 16.0–74.2 | 169 | 64 (56–70), 39–88 | 160 |
| Age at diagnosis, median (IQR), range (years) | 12.3 (7.9–20.6), 0.8–68.1 | 611 | 11.5 (7.9–17.0), 0.8–55.3 | 169 | 15 (8–23), 0–49 | 169 |
| Diabetes duration, median (IQR), range (years) | 18.5 (11.8–29.9), 3.8–72.6 | 611 | 17.2 (10.3–27.8), 5.0–64.8 | 169 | 45 (41–51), 35–72 | 160 |
| Sex (% female) | 59 | 611 | 59 | 169 | 45 | 175 |
| Blood pressure (mm Hg) |  |  |  | 163 |  | 155 |
| Systolic | 128 (13), 90–183 | 588 | 126 (12), 90–163 |  | 137 (18), 64–218 |  |
| Diastolic | 72 (8), 45–102 | 588 | 71 (8), 52–101 |  | 70 (10), 41–99 |  |
| Height (cm) | 175 (9), 145–200 | 610 | 174 (9), 156–193 | 169 | 175 (9), 146–203 | 160 |
| Weight (kg) | 79 (13), 52–123 | 592 | 78 (12), 54–113 | 166 | 81 (16), 38–134 | 161 |
| BMI (kg/m<sup>2</sup>) | 26 (4), 18–40 | 592 | 26 (4), 19–37 | 166 | 27 (4), 15–40 | 159 |
| Total daily insulin dose, median (IQR), range (U/day) | 50 (40–64), 15–179 | 598 | 52 (42–64), 15–130 | 165 | 30 (18–45), 8–124 | 114 |
| HbA1c |  | 609 |  | 169 |  | 112 |
| mmol/mol | 61 (13), 23–124 |  | 60 (12), 23–115 |  | 58 (9), 36–87 |  |
| % | 7.7 (1.1), 4.3–13.5 |  | 7.7 (1.1), 4.3–12.7 |  | 7.4 (0.8), 5.4–9.1 |  |
| Glucose (mmol/L) | 9.6 (3.9), 1.0–28.3 | 609 |  |  | - |  |
| C-peptide undetectable vs detectable (%) |  | 609 |  | 169 | - |  |
| Undetectable (<3.8 pmol/L) | 74 |  | 75 |  |  |  |
| Detectable (≥3.8 pmol/L) | 26 |  | 25 |  |  |  |
| C-peptide concentration in participants with detectable C-peptide, median (IQR), range (pmol/L) | 30 (10–110), 3.9–1439 | 155 | 31 (10–129), 4–741 | 43 | - |  |
| Insulin administration (%) |  | 610 |  | 169 |  | 150 |
| MDI | 39 |  | 33 |  | 73 |  |
| Insulin pump | 61 |  | 67 |  | 77 |  |
| Glucose monitoring method (%) |  | 610 |  | 169 |  | 120 |
| SMBG | 77 |  | 82 |  | 17 |  |
| rt-CGM | 19 |  | 14 |  | 47 |  |
| is-CGM | 4 |  | 4 |  | 36 |  |
| Total cholesterol (mmol/L) | 4.4 (0.8), 2.6–8.9 | 611 | 4.3 (0.8), 2.8–7.0 | 169 | 4.3 (0.9), 0.5–7.2 | 145 |
| HDL-cholesterol (mmol/L) | 1.7 (0.5), 0.6–3.6 | 611 | 1.7 (0.5), 0.9–3.6 | 169 | 1.7 (0.5), 0.3–3.1 | 146 |
| LDL-cholesterol, mmol/L | 2.7 (0.7), 0.8–5.8 | 611 | 2.6 (0.7), 1.3–5.2 | 169 | 2.4 (0.7), 0.9–5.4 | 147 |
| Triglycerides, median (IQR), range (mmol/L) | 0.9 (0.7–1.2), 0.2–5.4 | 606 | 0.8 (0.7–1.1), 0.3–4.7 | 164 | 1.0 (0.7), 0.4–6.4 | 147 |
| Apolipoprotein B (mmol/L) | 0.8 (0.2), 0.3–1.7 | 611 | 0.8 (0.2), 0.4–1.7 | 169 | - |  |
| Antihypertensive medication (%) | 21 | 595 | 14 | 168 | 34 | 159 |
| Lipid-lowering medication (%) | 23 | 596 | 19 | 168 | 26 | 159 |

<sup>a</sup> recruitment still ongoing
HbA1c, glycated haemoglobin; HDL, high density lipoprotein; IQR, interquartile range; is-CGM, intermittent scanning continuous glucose monitoring; LDL, low density lipoprotein; MMTT, mixed-meal tolerance test; rt-CGM, real-time continuous glucose monitoring; SD, standard deviation; SMBG, self-monitoring of blood glucose

**Figure 2.**
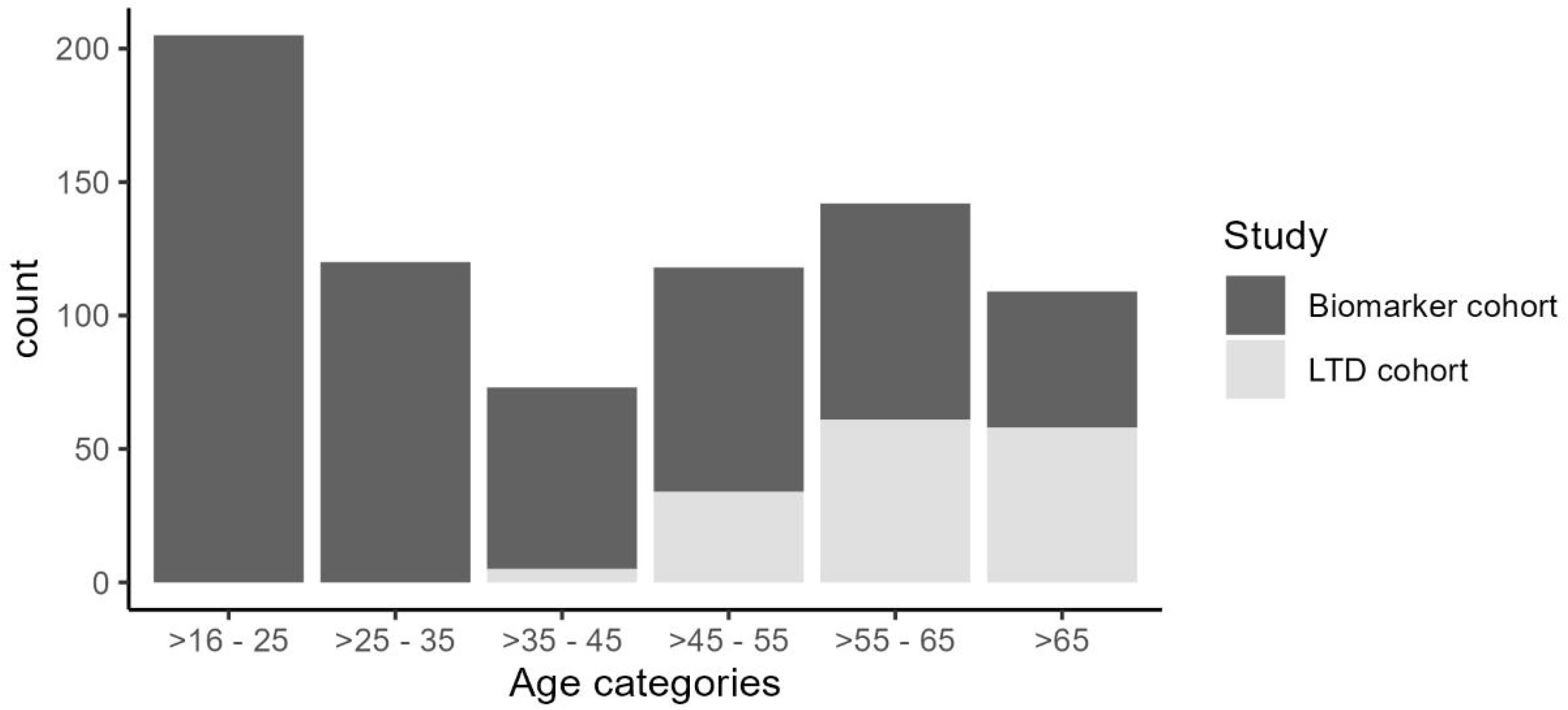
Distribution of age.

**Figure 3.**
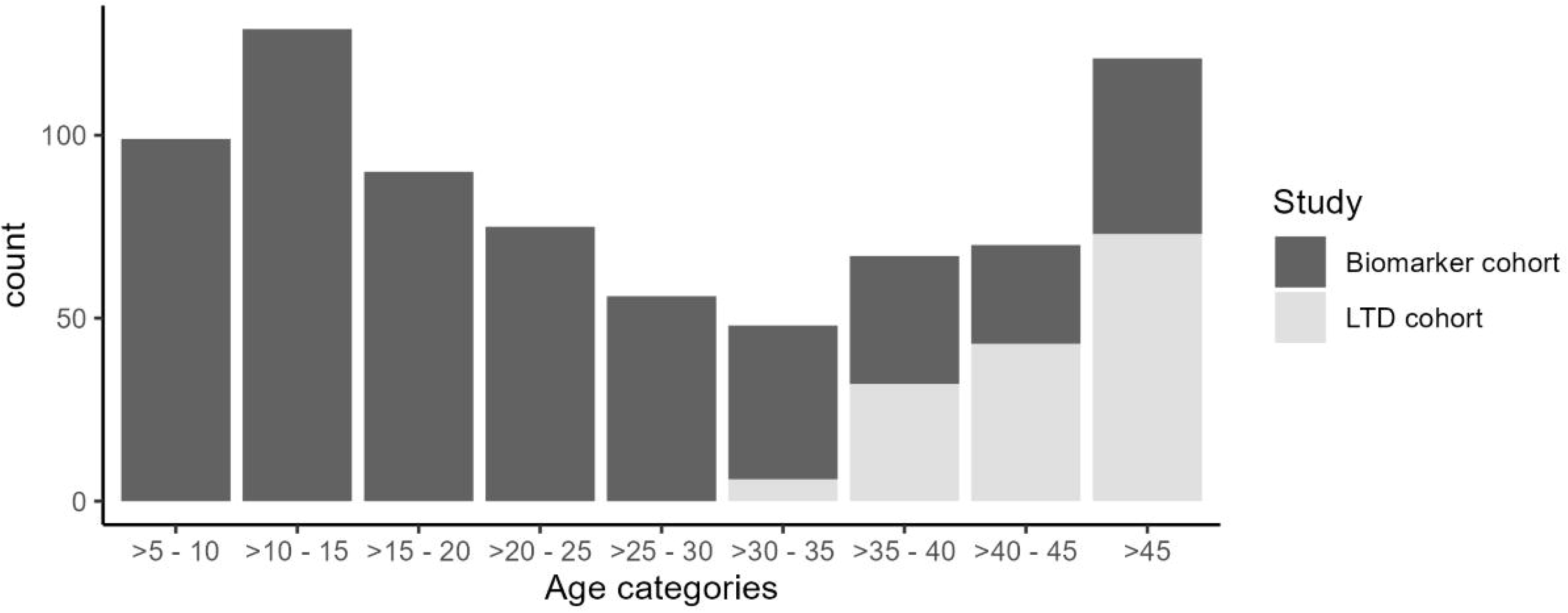
Distribution of diabetes duration.

The individuals in the LTD cohort had a median age and diabetes duration of 63 (56–70) and 45 (40– 50) years, respectively (also see figure 3). Almost half the participants were women with a mean BMI of 27 (4) kg/m². Mean (SD) HbA1c was 58 (9) mmol/mol (7.4 [0.8]%) with a median total daily dose of 30 (18–49) U/day. Around half the participants used an insulin pump, with 47% using rt-CGM and 36% using is-CGM. This is higher when compared with the Biomarker complete study cohort, likely because rt-CGM was reimbursed at the time this study started. Antihypertensive and lipid lowering drugs were used by 21% and 16% of the study participants, respectively.

### Follow-up information

Biomarker complete study cohort: After baseline recruitment of 611 participants (T0), 539 (88%) returned for a 1-year follow-up visit between May 2017 and March 2020 (T1), after a median follow-up time of 12.6 (11.9–13.5) months after T0 (range: 8.9–20.3). Two-year follow-up visits were completed by 460 participants (75%) between April 2018 and March 2021 (T2), after a median of 13.0 (11.7–13.6) months after T1 (range: 4.8–35.4). Reasons for loss to follow-up included, but were not restricted to, Covid-19 (40%), own request/no time (23%), diabetes treatment at other hospital (17%), pregnancy/breastfeeding (7%), illness/medication use/accident (5%), death (2%) and other reasons (6%).

MMTT-cohort: 169 participants (28%) underwent an MMTT between August 2016 and February 2019 2020, of whom 104 underwent a second MMTT (62% of the 169 participants who underwent an MMTT at baseline) between August 2017 and March 2019, after a median follow-up time of 12.4 (11.9–13.4) months (range: 8.6–25.3).

### Publications to date

#### 1. C-peptide assay validation and sample analysis

Samples from the cohort (only from one centre) have first been used to compare and verify two commercially available assays able to measure C-peptide in the picomolar range. Many C-peptide assays are commercially available for research and routine use. ^35^ We compared the ultrasensitive Mercodia enzyme-linked immunosorbent C-peptide assay (ELISA) with the Beckman immunoradiometric assay (IRMA) for C-peptide, assessing reproducibility (coefficient of variation [CV]), limit of blank (LoB), limit of detection (LoD) and limit of quantitation (LoQ). ^20^ Because only the IRMA met the specifications claimed by the manufacturer, providing the lowest threshold for quantification of serum C-peptide, we chose this assay for our C-peptide analyses. ^20^

#### 2. Residual C-peptide production and impaired awareness of hypoglycaemia (IAH)

In a following analysis of the longitudinal cohort we assessed the association between fasting serum C-peptide levels and the presence of impaired awareness of hypoglycaemia (IAH) in people with T1D.^19^ Clinical data and fasting serum C-peptide concentrations were related to the presence or absence of IAH, which was evaluated using the validated Dutch version of the Clarke questionnaire. Using a multivariable logistic regression model, associations between C-peptide and other clinical variables with IAH were investigated. We reported that residual C-peptide secretion was associated with lower risk of IAH and that higher BMI, the presence of microvascular complications and higher age at diabetes onset were independent risk factors for IAH in people with T1D.

#### 3. Fasting versus stimulated residual C-peptide production

About 25% of PWDs still showed some residual C-peptide production (Table 3). One of our analyses focussed on the stability of residual C-peptide production over time. ^21^ Because the MMTT procedure constitutes a burden to PWDs, an easier way to reliably assess residual C-peptide secretion is required, e.g. a simplified MMTT testing only 1 timepoint. To assess if fasting residual C-peptide production or residual C-peptide production at 90 minutes or 120 minutes after a MMTT may be sufficient to identify residual C-peptide production, we compared these variables with the C-peptide area under the curve (AUC) of the complete MMTTs. About 10% of participants who did not show fasting residual C-peptide production, did still show meal-stimulated residual C-peptide production. The 90 minutes and 120 minutes MMTT timepoints showed good concordance with the MMTT total AUC. Overall, there was a decrease of C-peptide at 1-year follow-up. ^21^

### Strengths and limitations

With regard to the prospective part of the study, the main strengths of this cohort are the prospective nature of the measurements and the collection of biosamples from participants with established T1D, whereas most longitudinal studies collect biosamples in the context of the early phases of T1D. To test stability of C-peptide assessments, a subgroup of participants underwent an MMTT at 2 time points. The clinical and biochemical data can be linked to the results from ancillary genetic studies on DNA and mRNA and a large pool of data and samples is available for collaborative projects. For both cohorts detailed clinical data are available of all participants and for a large subgroup of PWDs’ historical glucose and HbA1c data are also available. Furthermore, PWDs were recruited from multiple clinics, including non-academic/-specialised centres, representing a "real world" population.

This study also has some limitations. First, although this was a longitudinal study, participants were followed for only 2 years, which is relatively short. Second, this cohort may not be representative for the general Dutch population of PWDs, considering the high percentage of mainly younger participants using technology. It is likely that these participants feel more engaged in looking for solutions for T1D (management), potentially introducing participation bias. Third, the inclusion age of ≥16 years old prevents extrapolation of the data to younger cohorts, because the disease process in younger patients is different and more rapid at onset, resulting in less C-peptide reserve causing different disease processes later in the course of the disease. Fourth, in this study cellular immunity was not assessed so it is not possible to investigate possible associations between beta-cell function and beta-cell autoimmunity. Fifth, although data and biosamples were collected for more than 600 participants, this number may be too low for detailed analyses in specific PWD subgroups (e.g. insulin delivery modality, different treating centres and therapies). Finally, there was a relatively high dropout rate of 25% after 2 years, largely influenced by the Covid-19 outbreak.

### Next steps and call for further use of data and samples and collaborative efforts

Diabetes research on the intersection of immunology and metabolism is a field that is developing in a fast pace.^36^ While research is increasingly focusing on the role of the innate immune system in the earliest steps of the disease and its sequelae, ^37 38^ our main goal was to focus on heterogeneity in hormonal, immune/inflammatory/metabolic markers and insulin resistance in PWDs. With regard to hormones, the samples have already been analysed for fasting and stimulated C-peptide. Future biochemical analyses will be aimed at determining fasting and stimulated levels of additional relevant hormones such as glucagon, GLP-1, GLP-2, leptin, growth hormone and proinsulin. With respect to inflammatory markers in established T1D we plan to assess if innate immune markers (e.g. CRP, fibrinogen, complement component C3) or specific immune markers like cytokines (e.g. Il-6, Il-10, TNF-alpha) and adipokines (e.g. chemerin, CCL2, adiponectin) are associated with glucose metrics and residual beta-cell function (as measured by residual C-peptide production). In terms of glycaemic outcomes, HbA1c levels of participants were collected during the study. In addition, for participants from one centre (Diabeter Netherlands) automatically retrieved glucose (sensor) data since 2012 are available. These sensor data can be used for analysis of glycaemic variability parameters (e.g. time in targeted glucose [TIR], coefficient of variation [CV], mean amplitude of glycaemic excursions [MAGE] etc.), which can also be linked to the other data.

The innate immune system is able to sense metabolic stress induced by factors such as nutritional components and changes in the intestinal microbiota, refocusing the problem of diabetes to other organs, such as the liver and the gut. ^39^ We aim to explore some of the involved pathways in an integrated systems biology approach to assess clinical heterogeneity and to improve clinical phenotyping in T1D. While metabolic biomarkers associated with the early pathogenesis of T1D are established,^40^ there is a need for similar markers that exist beyond the clinical onset period. Currently, samples from the Biomarker cohorts are undergoing genetic, metabolomic and proteomic analyses. These data can then be linked to the hormonal data. We also plan to estimate additional features of insulin resistance based on blood pressure, insulin dose per kg lean body mass, markers of lipid metabolism (fasting free fatty acids, triglycerides and HDL-cholesterol), liver enzymes and adipokines (IL6, TNF-alpha, adiponectin).

In addition, new markers such as microRNA (miRNAs) have been identified as promising markers of disease in T1D, both in the disease process leading to beta cell destruction and in the subsequent development of complications. Circulating microRNAs and exosomes (EXOs: miRNA containing vesicles) are novel biomarker candidates, showing characteristic changes in blood levels in samples from PWDs.^41–45^ Also, it has been shown that miRNAs are transported within HDL and this way can reach target tissues to influence gene expression there. ^46^

All these data can be interlinked with data like residual beta-cell function (as measured by C-peptide levels), anthropometric and clinical parameters (e.g. [changes in] weight, length, glycaemic outcomes [HbA1c, glucose levels], insulin dose), and beta-cell-directed autoimmunity (e.g. [persistence of] autoantibodies, autoimmune comorbidities).

We welcome collaboration with other research groups interested in the data/samples of this cohort. Researchers can visit the Diabeter Research website (https://www.diabeterresearch.com/biomarker-study/) for additional information and can contact us at.

## Data Availability

All data produced in the present study are available upon reasonable request to the authors

## Acknowledgments

Establishing these cohorts and performing research would not have been possible without the interest, participation and support from all participating people with diabetes as well as the staff in all participating centres.

## Contributors

The Biomarker project was a Strategic Research Agreement (SRA) initially developed by HA, HV and BHRW with JDRF, later extended with the Diabetesfonds LTD cohort. HJA and BHRW conceived the present manuscript. MdV, HJA, BHRW and RDMV established the cohorts. PD, RDMV, CEV and EB conducted the data analysis. PD, DM and RDMV drafted the manuscript. All authors critically revised the manuscript and approved the final version of the manuscript. PD prepared the final version for the submission.

## Ethics approval Ethics approval

This study was approved by the Medical Ethics Review Board of the UMCG (METC 2015/493 and METC 2017/412). All participants gave written informed consent.

## Provenance and peer review

Not commissioned, externally peer reviewed.

## Data availability statement

Data are available upon request. Researchers can obtain additional information or submit a request and proposal for data and sample use to the Biomarker Steering Committee. Proposals are subject to a final decision by the Biomarker Steering committee.

